# Immunogenicity of the BA.1 and BA.4/BA.5 SARS-CoV-2 Bivalent Boosts: Preliminary Results from the COVAIL Randomized Clinical Trial

**DOI:** 10.1101/2023.01.31.23285306

**Authors:** Angela R. Branche, Nadine G. Rouphael, Cecilia Losada, Lindsey R. Baden, Evan J. Anderson, Anne F. Luetkemeyer, David J. Diemert, Patricia L. Winokur, Rachel M. Presti, Angelica C. Kottkamp, Ann R. Falsey, Sharon E. Frey, Richard Rupp, Martín Bäcker, Richard M. Novak, Emmanuel B. Walter, Lisa A. Jackson, Susan J. Little, Lilly C. Immergluck, Siham M. Mahgoub, Jennifer A. Whitaker, Tara M. Babu, Paul A. Goepfert, Dahlene N. Fusco, Robert L. Atmar, Christine M. Posavad, Antonia Netzl, Derek J. Smith, Kalyani Telu, Jinjian Mu, Mat Makowski, Mamodikoe K. Makhene, Sonja Crandon, David C. Montefiori, Paul C. Roberts, John H. Beigel

## Abstract

In a randomized clinical trial, we compare early neutralizing antibody responses after boosting with bivalent SARS-CoV-2 mRNA vaccines based on either BA.1 or BA.4/BA.5 Omicron spike protein combined with wildtype spike. Responses against SARS-CoV-2 variants exhibited the greatest reduction in titers against currently circulating Omicron subvariants for both bivalent vaccines.

## INTRODUCTION

The emergence of Omicron subvariants and waning immunity led to the authorization in various countries of SARS-CoV-2 bivalent vaccines that combine wildtype (ancestral) spike and either Omicron BA.1 or BA.4/BA.5 spike.^1,2^ Currently, the predominant circulating SARS-CoV-2 Omicron subvariants contain key mutations in the spike protein receptor binding domain (e.g., R346T) that enhance viral escape from neutralizing antibodies. The Coronavirus Variant Immunologic Landscape Trial (COVAIL) is an adaptive phase 2, open-label, randomized clinical trial assessing the immunogenicity of variant-containing SARS-CoV-2 vaccines, from different platforms, in previously vaccinated adults. Here we describe results from COVAIL Stage 4, where participants were randomized to a second boost with either the bivalent Pfizer/BioNTech BNT162b2 Wildtype/Omicron BA.1 vaccine or the Wildtype /Omicron BA.4/BA.5 vaccine to determine their ability to produce antibodies that neutralize past and contemporaneous SARS-CoV-2 variants.

## METHODS

### Study Design and Eligibility Criteria

This phase 2 open-label, randomized, clinical trial was performed at US sites (Supplemental Table 1), enrolling all participants in October 2022. Eligible persons were healthy adults between the ages of 18 to 49 years of age (with or without prior SARS-CoV-2 infection) who had received a primary series and a single boost with an approved or emergency use authorized wildtype COVID-19 vaccine (Supplemental Table 2) confirmed by a review of their vaccination card. The most recent vaccination and prior infection, if applicable, must have occurred at least 16 weeks prior to randomization. Full eligibility criteria are described at clinicaltrials.gov (NCT 05289037).

After providing written informed consent, participants underwent screening, including confirmation of COVID-19 vaccination, medical history, a targeted physical examination, and a urine pregnancy test (if indicated). Eligible participants were randomly assigned to one of two vaccines in a 1:1 ratio and immunogenicity samples were collected pre-vaccination (Day 1) and after vaccination on Days 15 and 29, and 3, 6, 9 and 12 months. Intercurrent SARS-CoV-2 infections were collected by passive surveillance. The trial was reviewed and approved by a central institutional review board and overseen by an independent Data and Safety Monitoring Board. The trial was sponsored and funded by the National Institutes of Health (NIH).

### Trial vaccine

The bivalent Pfizer/BioNTech BNT162b2 Wildtype /Omicron BA.1 and Wildtype /Omicron BA.4/BA.5 vaccines were provided by Pfizer BioNTech (total amount of 30 mcg mRNA per vaccine; 15 mcg for each strain). The vaccine candidates are manufactured similarly to their corresponding authorized or approved vaccines.

### Study outcomes

The primary objective was to evaluate humoral immune responses of candidate SARS-CoV-2 variant vaccines. The secondary objective was to evaluate the safety of candidate SARS-CoV-2 variant vaccines assessed by solicited injection site and systemic adverse events (AEs), which were collected for 7 days after vaccination; unsolicited AEs through Day 29; and serious adverse events (SAEs), new-onset chronic medical conditions (NOCMCs), adverse events of special interest (AESIs), AEs leading to withdrawal, and medically attended adverse events (MAAEs) through the duration of the trial. Immunologic and safety data are currently available through Day 29.

### Immunogenicity assays

SARS-CoV-2 neutralization titers, expressed as the serum inhibitory dilution required for 50% neutralization (ID_50_), were assessed at baseline and at Days 15 and 29, as described previously, using pseudotyped lentiviruses^3,4^ presenting SARS-CoV-2 spike mutations for different strains. All samples (101 per vaccine arm) were tested in a commercial lab (Monogram Biosciences, CA) for the following variants: the D614G (Wuhan-1 containing a single D614G spike mutation), B.1.617.2, B.1.351, B.1.1.529 (Omicron BA.1) and Omicron BA.4/BA.5. Omicron BQ.1.1 and Omicron XBB.1 neutralization titers were assessed on a random subset of 25 samples per vaccine arm, distributed roughly equally between previously infected and uninfected participants in the Montefiori Lab at Duke University. Electrochemiluminescence immunoassays (ELECSYS) were used for the detection of anti-nucleocapsid (N) (N-ELECSYS; Elecsys Anti-SARS-CoV-2 N, Roche, Indianapolis) at baseline.^5^

### Statistical analysis of Immunogenicity Endpoints

The primary objective of this study is to evaluate the magnitude, breadth, and durability of SARS-CoV-2 specific immune responses measured by geometric mean antibody titers (GMT) in serum samples with associated 95% confidence intervals (CI). No formal hypothesis tests were planned. The geometric mean fold rise (GMFR) is calculated as the geometric mean of titers at a timepoint divided by titers at Day 1. The geometric mean ratio to D614G (GMR_D614G_) is the ratio of the geometric mean titers for a variant of concern to titers against D614G. Seropositivity rate is calculated as the proportion of participants with titers above the lower limit of detection (LLOD). 95% CI for GMT, GMFR, and GMR_D614G_ are calculated using the Student’s t-distribution and the 95% CI is calculated using the Clopper-Pearson binomial method. For analysis, participants were defined as previously infected by self-report of a confirmed positive antigen or PCR test or by a positive anti-nucleocapsid (N) antibody test at enrollment. Participants with COVID-19 occurring between vaccination and a pre-specified immunogenicity timepoint were excluded from the immunogenicity analyses at all timepoints post infection.

## RESULTS

### Study Population

Two hundred two previously vaccinated and boosted participants were enrolled between October 4 – 28, 2022 and received either the bivalent Pfizer/BioNTech BNT162b2 Wildtype /Omicron BA.1 (n=101) or Wildtype /Omicron BA.4/BA.5 vaccines (n=101). Baseline characteristics were similar between the two study arms (Supplemental Table 3). Median age was 31 years (range: 18-49). The majority of participants (93% per arm) had received an mRNA-based primary series and boost vaccine. At enrollment, 77% were defined as previously infected by anti-N antibody seropositivity at baseline and/or by self-reported positive SARS-CoV-2 PCR or antigen testing (Supplemental Table 3). Median duration (range) between study vaccination and the last previous vaccination or infection was 293 (112-585) days.

### Safety

Solicited local and systemic AEs after vaccination were similar to other booster trials^6^ and did not differ between arms (94% for the Wildtype/Omicron BA.1 arm and 92% for the Wildtype/Omicron BA.4/BA.5 arm). The most frequently reported solicited local AE was injection-site pain (80%). The most common solicited systemic AEs were fatigue (68%) and myalgia (53%). Most solicited AEs were mild to moderate; only 1% of local AEs (induration/swelling) and 3% of systemic AEs (predominantly headache and fatigue in addition to fever, arthralgia, myalgia) in 7 participants were graded as severe. There were no AESI, SAEs or AEs leading to withdrawal from the study at the time of interim analysis. (Supplemental Figures 1 and 2 and Tables 10-12)

### Neutralizing Antibody Responses

All participants were seropositive against all variants after the boost, with titers peaking at Day 15 for all variants except D614G, which peaked at Day 29 (Figure 1). At Day 15, ID_50_ GMTs in the Wildtype/Omicron BA.1 arm were numerically similar (with overlapping confidence intervals) to corresponding titers in the Wildtype/Omicron BA.4/BA.5 arm for D614G (ID_50_ 27,000 vs. 34,109), BA.1 (ID_50_ 6,506 vs. 6,603), B.1.351 (ID_50_ 15,183 vs. 19,265) andB.1.617.2 (ID_50_ 14,362 vs. 18,332).However, Day 15 titers against Omicron BA.4/BA.5 were >1.5 higher with the Wildtype/Omicron BA.4/BA.5 (GMT_BA.4/BA.5_ = 5,939) compared to titers after vaccination with the Wildtype/Omicron BA.1 vaccine (GMT_BA.4/BA.5_ = 3,546) (Figure 1, Supplemental Tables 4-7). Similar findings were observed at Day 29. Titers from participants without a history of prior infection were lower (Supplemental Tables 6 and 7) than those with hybrid immunity (Supplemental Tables 4 and 5).

**FIGURE 1.**
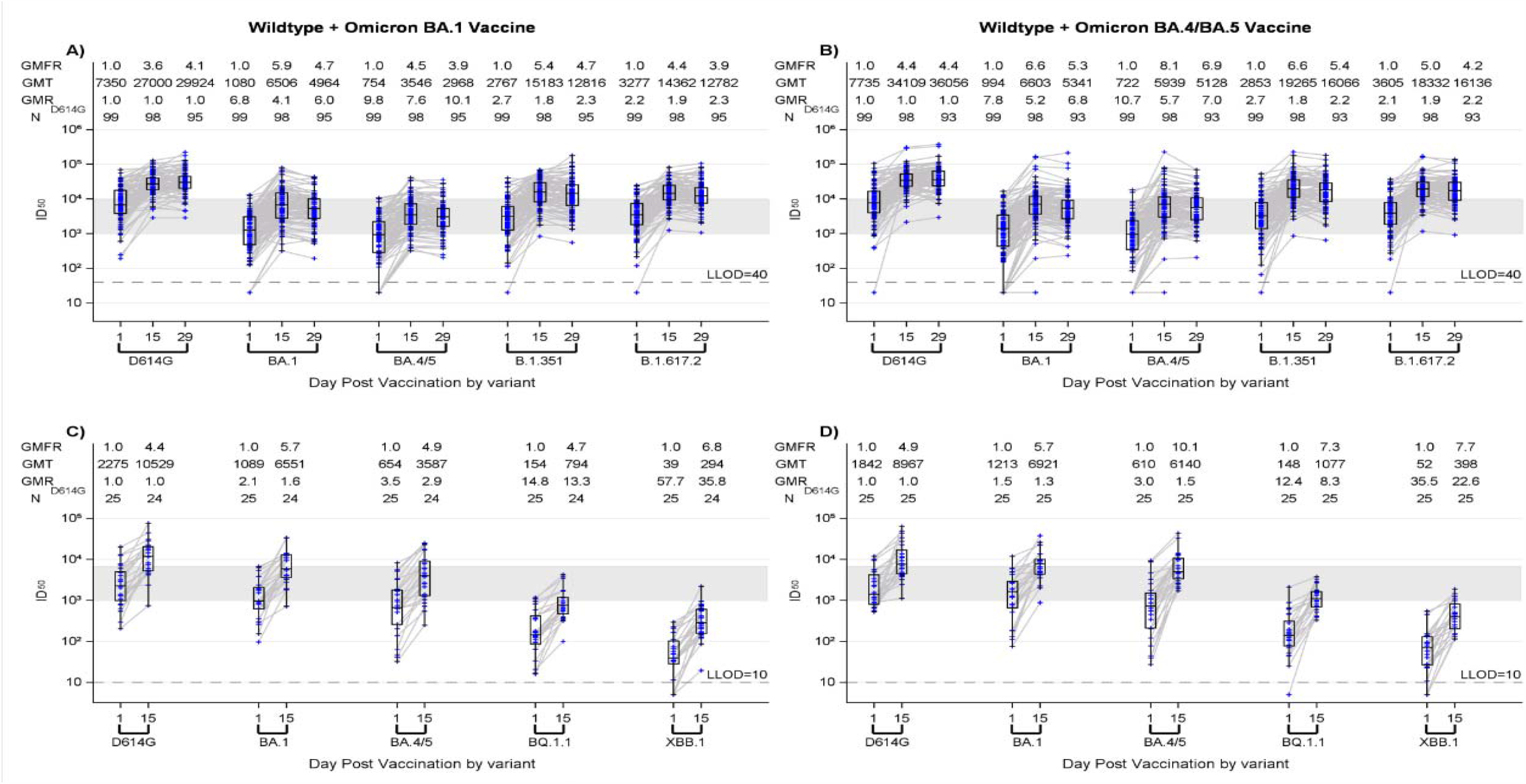
Pseudovirus Neutralization ID_50_ Titers by Timepoint (baseline, Day 15 and Day 29) and Variant before and after vaccination with 30 mcg of Pfizer/BioNTech BNT162b2 Wildtype/Omicron BA.1 (A and C) or 30 mcg Pfizer/BioNTech BNT162b2 Wildtype/Omicron BA.4/BA.5 (B and D). Panels A and B show results from the Monogram lab for each vaccine candidate against D614G, Omicron BA.1 [B.1.1.529], BA.4/BA.5, B1.351 [Beta], B.1.617.2 [Delta] at baseline, Days 15 and 29 post vaccination. Panels C and D show results from the Duke University Montefiori lab for each vaccine candidate against D614G, Omicron BA.1 [B.1.1.529], BA.4/BA.5, BQ.1.1 and XBB.1 at baseline and Day 15 post vaccination. Boxes with horizontal bars denote interquartile range (IQR) and median ID_50_, respectively. Whisker denotes 95% confidence interval. LLOD, lower limit of detection of the assay. GMT, geometric mean titer. GMFR, geometric mean fold rise from baseline

Titers against all Omicron subvariants were lower than against D614G; the lowest titers were observed against XBB.1 (Figure 1). Notably, titers against BQ.1.1 and XBB.1 were similar between the two arms (with overlapping confidence intervals). Titers against BQ.1.1 and XBB.1 were 8-22 times and 13-35 times lower than against BA.1 and D614G, respectively, with the Wildtype/Omicron BA.1 vaccine. Titers against BQ.1.1 and XBB.1 were 4-12 times and 8-22 times lower than against BA.4/BA.5 and D614G, respectively, with the Wildtype/Omicron BA.4/BA.5 vaccine (Figure 1, Supplemental Tables 8-9).

## DISCUSSION

Our study is the only randomized trial to date to report results from a head-to-head comparison of the two mRNA Wildtype/Omicron (BA.1 or BA.4/BA.5) bivalent vaccines currently authorized worldwide as a boost in individuals previously immunized with a first generation COVID-19 vaccine series. Our early immunogenicity results demonstrate better neutralization against BA.4/BA.5 with the Wildtype/Omicron BA.4/BA.5 vaccine. However, there was increasing neutralization escape with the late 2022 Omicron subvariants (BQ.1.1 and XBB.1). This escape is similar between the two bivalent vaccines as demonstrated by numerically similar GMTs with overlapping confidence intervals, even though BA.1 and BA.4/BA.5 spike sequences are known to have different mutations in the receptor binding domain.^7^

Though modest serologic advantages to Omicron BA.1 and BA.4/BA.5 have been previously reported with bivalent compared to wildtype vaccines,^8^ we do not currently have precise immune correlates of protection for emerging variants. Moreover, we did not evaluate the immunogenicity of the wildtype vaccine since this vaccine is no longer recommended as a boost in the US. We conducted passive surveillance for SARS-CoV-2 intercurrent infections and some cases could have been missed that could confound our immunogenicity results. These early preliminary results also do not address durability and the conclusions about protection are limited by the small sample size. Finally, serologic data from timepoints up to 1 year after vaccination, and cellular responses, which are known to influence disease severity, are also pending.

However, our findings highlight ongoing concern that the breadth of antibody response from current updated vaccines is not optimal for the pace of virus evolution. Consequently, while early vaccine effectiveness (VE) data with bivalent vaccines have emerged,^9,10^ continuous surveillance is crucial to assess for potential VE waning.

## Data Availability

All data produced in the present study are available upon reasonable request to the authors.

## Acknowledgments

We would like to thank all participants who contributed to the study and the NIAID SARS-CoV-2 Assessment of Viral Evolution (SAVE) program team for their consultation regarding the study arm design and variant vaccine selection.

## Conflicts of Interests

− **EJA** has received grants from Pfizer, Moderna, Janssen, GSK, Sanofi, Micron, and Regeneron through institution as well as consulting fees from Pfizer, Janssen, Moderna, and Sanofi. EJA serves on safety/advisory boards for Sanofi, ACI Clinical/WCG and Kentucky Bioscience, Inc.
− **ARB** has received research support from NIH-NIAID, grants from Pfizer, Cyanvac, and Merck as well as consulting fees from Janssen and GSK.
− **LRB** has received grants from Wellcome Trust, Gates Foundation, NIH/Harvard Medical School through institution. Serves as member of DSMB for NIH and AMDAC for FDA. Dr Baden is involved in HIV and SARS-CoV-2 vaccine clinical trials conducted in collaboration with the NIH, HIV Vaccine Trials Network (HVTN), Covid Vaccine Prevention Network (CoVPN), International AIDS Vaccine Initiative (IAVI), Crucell/Janssen, Moderna, Military HIV Research Program (MHRP), the Gates Foundation, and Harvard Medical School.
− **DJD** has received a contract from Leidos Biomedical research to conduct the clinical trial through institution.
− **ARF** has received grants from Janssen, Pfizer, Merck, BioFire Diagnostics, and CyanVac through institution, consultant fees from Arrowhead and Icosavax, and honoraria as a speaker from Moderna and GlaxoSmithKline. ARF also serves on safety/advisory boards for Novavax and received travel/meeting support from GlaxoSmithKline.
− **SEF** has received f unding from Leidos to Saint Louis University to conduct Protocol DMID22-0004.
− **DNF** has as a contract from CDC and is the site PI for clinical trials from Gilead, Regeneron and MetroBiotech LLC. She is the PI on one investigator-initiated award from Gilead and the co-PI on another investigator initiated award from Gilead. DNF served on an HBV Advisory board for Gilead in 2021 and received payment for expert testimony not related to COVID in 2022.
− **PAG** has received funding for COVAIL clinical trial. PAG has also received consulting fees from Janssen Vaccines.
− **LCI** has received support for the present manuscript from NIH-NIAID/DMID, Moderna, Pfizer, and Sanofi. LCI has also received grants from GSK, Merck, Sharpe & Dohme Corp, CDC, Novavax, AHRQ, and NIH/NLM/NIMHD as well as consulting fees from Moderna, CDC, and Pediatric Emergency Medicine Associates, LLC. LCI has received honoraria as a speaker from American Academy of Pediatrics, Rockefeller University, and American Academy of Pediatrics-Georgia Chapter. LCI Serves on Data Safety Monitoring for NIH-Phase 2 Vaccine Trial for Monkeypox, Moderna Scientific Advisory Board-North America, and CoVID-19 Task Force, Georgia. LCI has a leadership role in the Pediatric Infectious Disease Society and serves as board member on the Emory University-Pediatric and Reproductive Environmental Health Scholars-Southeastern, the Center for Spatial Analytics of the Georgia Institute of Technology, and the American Academy of Pediatrics (Executive Board for Section on Infectious Diseases). LCI has received travel/meeting support from the American Academy of Pediatrics and Moderna.
− **LAJ** has received funding from NIH for support for this study, funding from Pfizer to support a clinical trial and contract funding for research support from the CDC and the NIH, all through institution. LAJ also reports unpaid participation on Data Safety Monitoring Boards for NIH funded clinical trials.
− **SJL** has received NIH grants through institution.
− **AFL** has received grants from Merck, Gilead and, Viiv through institution as well as consulting fees from Vir Biotechnology. AFL has also received travel support from Merck to attend a required investigator meeting, testing kits and supplies to support research study from Hologic, and medication donated by Mayne Pharma to support research study.
− **MM** has received funding from Division of Microbiology and Infectious Diseases for contract # 75N93021C00012.
− **DCM** has received funding from NIH/75N93019C00050-21A: CIVICS A-Option 21A-DMID Trials of COVID-19 Vaccines.
− **JM** has received funding from Division of Microbiology and Infectious Diseases, contract # 75N93021C00012.
− **AN** has received support from NIH-NIAID, CEIRR (Centers of Excellence for Influenza Research and Response) and Gates Cambridge Trust as well as grants from NIH-NIAID R01.
− **RMN** has received grants from Moderna and Janssen and travel/meeting support from Moderna.
− **CMP** has received funding from NIAID UM1AI148684.
− **RMP** has received funding from NIH DMID COVAIL as well as grants from Janssen, Moderna and NIH through institution.
− **NGR** has received research grants from Pfizer, Merck, Sanofi, Quidel and Lilly through institution, consulting fees from Krog, honoraria as speaker for Virology education, and travel support from Sanofi. NGR serves on safety committees for ICON and EMMES and is a member of the Moderna Advisory board.
− **DJS** has received support from NIH-NIAID CEIRR, grants from NIH-NIAID R01, and travel support from NIH-NIAID CEIRR for NIH-related meetings.
− **KT** has received funding from Division of Microbiology and Infectious Diseases contract # 75N93021C00012.
− **EBW** has received funding from Leidos Biomedical Research AGREEMENT NO. 22CTA-DM0009 as well as grants from Pfizer, Moderna, Sequiris, Clinetic, and Najit Technologies, with payments made to institution. EBW has also received honoraria as a speaker from College of Diplomates of the American Board of Pediatric Dentistry, consulting fees from Iliad Biotechnologies, and travel/meeting support from the American Academy of Pediatrics. EBW serves as member of Vaxcyte Scientific Advisory board.
− **PLW** has received subcontract funding from NIH for this study as well as NIH grant funding and contract funding from Pfizer through University of Iowa. PLW has also received consulting fees from Pfizer and serves on safety/advisory board for Emmes Corporation.

<u>The following group has no conflicts to declare:</u>

- **RLA, TMB, SMM, MB, MKM, JHB**, **SC, ACK, CL, PCR, RR, JAW**.

