## Supplementary Appendix for "Immunogenicity of the BA.1 and BA.4/BA.5 SARS-CoV-2 Bivalent Boosts: Preliminary Results from the COVAIL Randomized Clinical Trial"

**SUPPLEMENTARY MATERIALS**

COVAIL Manuscript Study Members

**University of Rochester VTEU, Rochester, NY**

Angela R. Branche, MD, Ann R. Falsey, MD, Edward Walsh, MD, Patrick Kingsley, BS, Arthur Zemanek, BSN, MS Katherine Elena, BSN, Spencer Obrecht, BSN, Ian Shannon, BSN, Amy Kaychalo, BS, MS, Erin Nowicki, Sharon Moorehead, Kari Steinmetz, BA, Doreen Francis, RN, Tanya Smith, BS, William Hamilton, BS, Jeanne Holden-Wiltse, MPH, MBA, Christopher Lane, MS, Michael Peasley, BS, Samuel Diehl, BS, Kyle Richards, PharmD, Stephen Bean, PharmD, Nicole Dornbush, PharmD, Carol Cole, PharmD

**Emory University Hope Clinic, Decatur, GA**

Nadine G. Rouphael, MD; Cecilia Losada, MD; Daniel S. Graciaa, MD; Hady Samaha, MD; Cassie Grimsley Ackerley, MD; Kristen E. Unterberger, PA; Amy Anderson, BSN; Mary Atha, ACNP; Kareem Bechnak, BSN; Sarah Bechnak, BSN; Mary Bower, BSN; Laura Clegg, RN; Matthew Collins, MD, PhD; Francine Dyer, RN; Srilatha Edupuganti, MD; Rebecca Fineman, BS; Tigisty Girmay, MSN; Rebecca Gonzalez, PharmD; Natalie Gray, BS; Evan Gutter, MPH; Lisa Harewood; Chris Huerta, MSc; Brandi Johnson, BS; Lauren Johnson, MPH; Colleen Kelley, MD; Alexandra Koumanelis, BA; Deborah Laryea, BSN; Hollie Macenczak, BSN; Nour Makkaoui, MD; Michele McCullough, MPH; Tuong-Vy Ngo, PharmD; Eileen Osinski, BS; Julia Paine, BS; Bernadine Panganiban, BS; Rose Pope, RN; Paulina Rebolledo, MD; Susan Rogers, RPh; Erin Scherer, PhD; Veronica Smith, NP-C; Andre Stringer, BS; Jessica Traenkner, PA; Dongli Wang, BS; Alahna Watson, BA; Stacey Wheeler, RN; Jean Winter; Jianguo Xu, PhD

**Brigham and Women’s Hospital, Harvard Medical School, Boston, MA**

Lindsey R. Baden, MD; Amy C. Sherman, MD; Stephen R. Walsh, MD; Alexandra Tong, BS; Rebecca Rooks, BS; Jane A. Kleinjan, NP; Jon A. Gothing, NP; Andres A. Avila Paz, BA; Muneerah M. Aleissa, PharmD, MPH; Bethany Evans, BA, August Heithoff, BS; Natalie E. Izaguirre, MS; Hannah Jin, MPH; Urwah Kanwal, BS; Austin Kim, BS; Julia E. Klopfer, BS; Christina Montesano, BS; John Almeida, BA; Emily S. Koleske, BS; Hannah Levine, BS; Nicholas P. Morreale, BS; Omolola Ometoruwa, BS; Jun Bai Park Chang, BS; Anna F. Piermattei, BA; Djenane M. Pierre, BS; Megan Powell, BA; Kevin Zinchuk, PharmD; Stephanie Pickford, PharmD; Charles M. Kelly III, PharmD; Xiaofang Li, PhD; John Kupelian, BS; Kimberly Dufresne, BS; Xiaoguang Fan, MD, PhD; Xi Zhang, PhD; Esther Arbona-Haddad, MD; Jose Humberto Licona, MD

**Center for Childhood Infections and Vaccines (CCIV) of Children’s Healthcare of Atlanta and Emory University Department of Pediatrics, Atlanta, GA**

Evan J. Anderson, MD; Christina A. Rostad, MD; Satoshi Kamidani, MD PhD; Etza Peters, RN; Larry Anderson, MD; Julia Bartol; Leisa Bower RN; Natsuko Campbell RN; Lisa Harewood, Hui-Mien Hsiao, Laila Hussaini MPH, Inara Jooma, Gidget Kettle RN; Marcia Lewis RN; Wensheng Li; Cindy Lubbers RN; Lisa Macoy RN; Molly Morrison, Heather Nurse RN; Anna Siaw-Anim; Kathleen Stephens RN; Madeline Taylor; Ashley Tippett MPH; Lauren Nolan PA

**Zuckerberg San Francisco General, University of California San Francisco, San Francisco, CA**

Anne F. Luetkemeyer, MD; Chloe Harris, BA; Azquena Munoz Lopez, BS; Daniel Berrner; Dennis Dentoni-Lasofsky, MSN; John Dwyer, RN; Suzanne Hendler, BSN; Elvira Gomez, MPH; WeyLing Phuah, PharmD; Jaime Velasco, BA; Veronica Viar, MS

**George Washington Vaccine Research Unit, George Washington University, Washington D.C.**

David J. Diemert, MD; Elissa Malkin, DO; Jeffrey M. Bethony, PhD; Aimee Desrosiers, PA-C; Marc Siegel, MD; Nikita Schroll-McLaughlin, MS; Jonathan Manning, BA; Jane Ryu, MS; Hanna-Grace Rabanes, MPH; Khadija Khan, MPH; Laura Vasquez, MPH; Caroline Thoreson, PA-C; Larissa Scholte, PhD; Rafaela Thur, DVM; Peyton St. John, BS; Dorinne Mettle-Amuah, PharmD

**University of Iowa College of Medicine, Iowa City, IA**

Patricia L. Winokur, MD; Jeffery Meier, MD; Jack Stapleton, MD; Laura Stulken, PA; Theresa Hegmann, PA; Deb Pfab, RN; Elizabeth Morgan, RN; Susan Herman, RN; Angel Peguero, CMA; Michelle Rodenburg; Alfred J. Carr; Delilah Johnson

**Washington University School of Medicine, St. Louis, MO**

Rachel M. Presti, MD, PhD; Jane A. O’Halloran, MD, PhD; Michael Klebert, RN, PhD Ryley M. Thompson; Alem Haile; Kim Gray, NP; Chapelle Ayres; Delaney Carani, RN; Michael Royal; John Tran; Laura Blair; Anita Afghanzada; Natalie Schodl

**NYU VTEU Manhattan Research Clinic at NYU Grossman School of Medicine, New York, NY**

Angelica C. Kottkamp, MD; Tamia Davis, NP; Celia Engelson, NP; Vijaya Soma, MD; Abdulwahab Abdulai; Ashanay Allen; Natella Aronova, NP; Philip Aziz, PharmD; Emily Beato; Samuel Bliss, PharmD; Jacqueline Callahan, RN; Ellie Carmody, MD; Amanda Dontino, BS; Aimee Edwin, RN; Shelby Goins; Sarah Haiken; Ramin Herati, MD; Abdonnie Holder; Janice Hong; Trishala Karmacharya; Manpreet Kaur, PharmD; Hye-Youn Kim; Alexander McMeeking, MD; Mark Mulligan, MD; Wai Ng; Edward Nirenberg; Irma Noriega, NP; Samuel Nweke; Lalitha Parameswaran, MD; Levonne Phillip, MPH; Stephanie Rettig, MPH; Marie Samanovic-Golden, PhD; Madalyn Saporito; Pamela Suman; Meron Tasissa; Michael Tuen; Julia Wagner, MPH; James Wilson; Doris Wong, PharmD; Grace Yip, BS; Samantha Yip, RN; Heekoung Youn, RN; Lisa Zhao

**Saint Louis University, Center for Vaccine Development, St. Louis, MO**

Sharon E. Frey, MD; Getahun Abate, MD, PhD; Zacharoula Oikonomopoulou, MD; Daniel F. Hoft, PhD, MD; Irene Graham, MD; Azra Blazevic, DVM, MPH; Tamara Blevins, MS; Kathleen Chirco, BSN; Sabrina M. DiPiazza, BSN, MA; Stanley Doublin; Heather Hoertel Douds, MSNS, BSN; Carol G. Duane, PhD, RN; Eric Eggemeyer, BA; Linda M. Eggemeyer-Sharpe, BSN; Lauren Nicole Foreman, BSN; Sarah Louise George, MD; Geoffrey J. Gorse, MD; Michelle Harris, PharmD; Helay Hassas, PharmD; Rong Hou, MD; Ryan Clark Kerr, BSN; Kate Elizabeth Liefer, BSN; Melissa J. Loyet, RN; Lainey Mejia-Jauregui, BS; Keith Meyer, BS; Tracy Renee Montauk, BSN; Karla J. Mosby, RN; Amanda Nethington, BS; Huan Ning, MD; Nicole Purcell; Joan M. Siegner, BSN, MA; Janice M. Tennant, BSN, MPH; Mei Xia, PhD; Kiana Wilder, BA; Yinyi Yu, BS; Cassandra Nicole Zehenny, BSN

**University of Texas Medical Branch, Galveston, TX**

Richard Rupp, MD; Laura Porterfield, MD; Amber Stanford, PA-C; Robert Cox, RN; Kristin Pollock, RN; Diane Barrett, MS; Gerrianne Casey, RN; Amy McMahan, LVN; Cori Burkett, PA-C; Essie Cox

**NYU VTEU Long Island Research Clinic at NYU Long Island School of Medicine, Mineola, NY**

Martín Bäcker, MD, Kimberly Byrnes, RN, Asif Noor, MD, Andrew B. Fleming, MD, Sigridh A. Muñoz-Gómez, MD, Diana Badillo, MD, Steven E. Carsons, MD, Sajumon K. Joseph, FNP, Sarah J. Pastolero, RN, Sophie Danziger, Monica Benitez, Maung Aung, Louis Ragolia, PhD, Alicia Vasile, RPh, April Correll, RPh, Christopher Hall, Thomas Palaia, Miloni H Thakker, MD, Lavern Harvey, Lisa Zhao

**University of Illinois at Chicago-Project WISH, Chicago, IL**

Richard M. Novak, MD; Benjamin G. Ladner, MD; Andrea Wendrow, RPh; Jesica Herrick, MD; Alfredo J. Mena Lora, MD; Scott A. Borgetti, MD; Diana L Bahena, APRN; Regina Harden, BA; Renyce Powell;  David C. M. Chan, PharmD; Rebeca F. Gasari, PharmD; Michael Pacini, PharmD; Margarita M. Villarreal, CPhT; Rodrigo Reyes, ADN; Samuel M. Rene, MPH; Shannon M Whitted, BSN; Habiba Sultana, MBBS; Nanu Kunwar, BS; Tasmin Sultana, MBBS; Md R. Amin, PhD;  Mahmood Ghassemi, PhD,  Liam Morrissy, BS; Nia O'Neal, BS; Chasity Serrano, BS; Charlie Peterson, BA

**Duke Human Vaccine Institute, Duke University School of Medicine, Durham, NC**

Emmanuel B. Walter MD, MPH; Michael J. Smith MD, MSCE; M. Anthony Moody, MD; Kenneth E. Schmader, MD; Susan Doyle; Lynn S Harrington BSN; Lori Hendrickson BSN; Amy O’Berry MSN; Sherry Huber BSN; Janet Wootton RN, RSCN; Kelly Clark BA; Lani Banez; Stephanie Smith BA; Byron Hauser BS; Ally Odom BA; Emily Randolph BA; Krystina Yoder BA; Kathlene Chmielewski; Luis Ballon BA; Aubree Latorre; Breana Montgomery; Antony Tritz MS; Thad Gurley, MS; Margaret Pendzich

**Kaiser Permanente Washington Health Research Institute, Seattle, WA**

Lisa A. Jackson, MD, MPH; Maya Dunstan, MS, RN; Rebecca Lau, PharmD; Barbara Carste, MPH; Wesley A. Andersen, RPh, MHA, MA; Lee Barr, RN; Cassandra Bryant, BS; Joe Choe, BS; Lynn Gross, PA-C; Erika Kiniry, MPH; Bonnie Y Lam, PharmD; De Vona Lang; Stella Lee, BA; Paula J Lins, PA-C, MPH; Amy Mohelnitzky, PA-C; Marilyn Nguyen, BS; Matthew Nguyen, MPH; Melissa Resendiz Rivas, BA; Melissa Boothe Scheer, PA-C; Janice Suyehira, MD; Stacie Wellwood, LPN; Maryann K Woodford, PA-C

**Department of Medicine, Division of Infectious Diseases and Global Public Health, University of California San Diego, La Jolla, CA**

Susan J. Little, MD; Thomas C.S. Martin, MD; Nicole Carter, MPH; Steven Hendrickx, RN; Ajay Bharti, MD; Alyssa Phillips; Aurora Verduzco Gonzalez, NP; Cheryl Dullano; Chris Houston; Dawn Rosenblum, RN; DeeDee Pacheco; DeLys Brooks; Fang Wan; Helene Le, CPhiT; JC Alcantar; Jill Blumenthal, MD; Joseph Lencioni, MABMH; Kory Hess; Letty Muttera, PharmD; Marlene Arredondo; Megan Smyth; Megan Taylor; Melinda Stafford, PharmD; Michelle Orsburn, MD; Michelle Truong; Niamh Higgins, PharmD, MSc, AAHIVP; Nimish Patel, PharmD, PhD, AAHIVP; Rebecca Gonzalez; Vivian Maldonado

**Morehouse School of Medicine, Atlanta, GA**

Lilly C. Immergluck, MD; Erica Johnson, PhD; Austin Chan, MD; Fatima Ali, MPH; Sonja Jackson; Noor Mohamed, PharmD; LaKesha Tables, MD; Norberto Fas, MD; Kay Woodson, PharmD; Saadia Khizer, MD; Jacquelyn Ali, MSA; Abdullah Warsama; Eric Gaines; Sierra Jordan Thompson; Cristina Wilson; Trisha Parker, MPH; Xiting Lin; LaTeshia Thomas Seaton, APRN, Derrick Wilson

**Howard University College of Medicine, Howard University Hospital, Washington D.C.**

Siham M. Mahgoub, MD; Celia Maxwell, MD; Sarah Shami, PharmD; Edward Bauer, BS; Yuanxiu Chen, MD, PhD; Megan Ware-Pressley, MHA; Debra Ordor, RN; Linda Fletcher, RN; Emmanuel Baidoo, BS; David Jaspan, RPh, MBA; Adetokunbo Adedokun, PharmD, MPH, BCPS; Michelle Strobeck, BS; Michael A. Riga; Ashley Karen Bautista, BS

**Departments of Molecular Virology and Microbiology and Medicine, Baylor College of Medicine, Houston, TX**

Jennifer A. Whitaker, MD; Hana M. El Sahly, MD; Wendy A. Keitel, MD; C. Mary Healy, MD; Robert L. Atmar, MD; Pedro A. Piedra, MD; Jesus Banay; Kathy Bosworth; Janet Brown, RPh; Kayla Burrell; Jeremy Castro; Tykel Eddy; Marcena Eubanks; Cathy Faw, RPh; Rachel Froebe; Alix Halter, RN; Janey John, MSN, APRN, FNP-C; Chanei Henry, AAS; Vanessa Martinez; Carol Mundell, RN; Brandie Phillips, RN; Alicia Prevost-Barthe, RN; Connie Rangel, RN; Yolanda Rayford, MS; Yvette Rugeley; Maria Shlyapobersky; Tina Sierra; Elizabeth Silguero; Lisreina Toro; Dawn Turner, RN; Chianti Wade-Bowers, RN; Jessica Woods, RN

**Departments of Medicine, Epidemiology, and Laboratory Medicine & Pathology, University of Washington, Vaccines and Infectious Diseases Division, Fred Hutchinson Cancer Center, Seattle, WA**

Tara M. Babu, MD, MSCI; Anna Wald, MD, MPH; Taylor Krause, BA; Kirsten Hauge, MPH; Jina Taub, ARNP, Dana Varon, ARNP, Britt Murphy, ARNP, Morissa Pertik, PA-C, T. Nui Pholsena, ARNP, Alyssa Braun, BS, Mark Drummond, BS, Jessica Heimonen, MPH, Amy Link, BS, Lindsey McClellan, BS, Jessica Moreno, BS, Chloe Wilkens, BS, Matt Seymour, MPH, Lawrence Hemingway, BS, Jean Mernaugh, BS, Chris McClurkan, BS, Kerry Laing, PhD, Meredith Potochnic, PharmD, Joong Kim, PharmD, Bao-Chao Vo, PhT, Dil Singh, BS

**University of Alabama at Birmingham, Birmingham, AL**

Paul A. Goepfert, MD; Jenna Weber, RN; Savannah Spaulding, RN; Heather Logan, CRNP; Faye Heard; Foreamben Patel; Michelle Chambers

**Tulane University School of Medicine, New Orleans, LA**

Dahlene N. Fusco, MD; Arnaud C. Drouin, MD; Florice K. Numbi, MD; Hamada F. Rady, PhD; Crystal A. Ward, MSN; Quinn M. Powers, MS; William E. Casey, BS; Brian P. Logarbo, MD; Shae P. Williams, BS; Emily Callegari, MSN

**IDCRC Principal Investigators**

David S. Stephens, MD; Kathleen M. Neuzil, MD

**IDCRC Leadership Operations Center**

Monica M. Farley, MD; Jeanne Marrazzo, MD; Sidnee Paschal Young

**IDCRC Clinical Operations Unit**

Jeffery Lennox, MD; Robert L. Atmar, MD; Linda McNeil FHI360

**IDCRC Laboratory Operations Unit – Fred Hutchinson Cancer Center and University of Washington, Seattle, WA**

Christine M. Posavad, PhD; Megan A. Meagher, BS; Michael Stirewalt, MBA; John Hural, PhD; Weston Lawler, BA; Lexi Tanser, MA, Julie McElrath, MD, PhD; Mike Gale, PhD

**IDCRC Statistical and Data Science Unit**

Elizabeth Brown, PhD

**The Emmes Company, LLC, Rockville, MD**

Mat Makowski, PhD; Heather Hill, MS; Jim Albert, MS, Holly Baughman; Lisa McQuarrie, MS; Kalyani Telu, MS; Jinjian Mu, PhD

**Clinical Monitoring Research Program Directorate, Frederick National Laboratory for Cancer Research, Frederick, MD**

Teri C. Lewis, BS; Lisa A. Giebeig, MS; Theresa M. Engel, MFS.; Caleb J. Griffith, MPH; Wendi L. McDonald, BSN; Alissa E. Burkey, MS; Lisa B. Hoopengardner, MS; Jessica E. Linton, MS; Nikki L. Gettinger, MPH; Aroussiak Bowen; Beth R. Baseler, MS; Vanessa S. Eccard-Koons, MS; Charles W. R. Hofsommer, JD; Thomas C. Sova, JD; Gary A. Krauss

**Duke Laboratory, NC**

David C Montefiori, MD, Amanda Eaton, MBA, Francesca Suman, MS.

**Centre for Pathogen Evolution, Department of Zoology, University of Cambridge, Cambridge, UK**

Derek J Smith, PhD; Antonia Netzl; Samuel H Wilks, PhD; Sina Tureli, PhD; Ana Mosterín Höpping, PhD; Samuel Turner; Sarah James, MD; Poppy Roth

**Division of Microbiology and Infectious Diseases, National Institute of Allergy and Infectious Diseases, National Institutes of Health, Bethesda, MD**.

Marina Lee, PhD; Mamodikoe Makhene, MD; Mohamed Elsafy, MD; Rhonda Pikaart-Tautges, BS; Janice Arega, MS: Binh Hoang, RPh; Dan Curtin; Hyung Koo, BSN; Elisa Sindall, BSN; Aya Nakamura, RN, MS; Audria Crowder, BS; Guinevere Chun, RN, BSN, MSHS; Frank Kenny, PhD MPH; Seemi Patel, RHP, PharmD; Sonia Gales, MS; Ahsen Khan, JD; Walla Dempsey, PhD; Robert Jurao- RN, BSN; Sonja Crandon, BSN; Seema U. Nayak, MD; Marciela M DeGrace, PhD; Diane J Post, PhD; Paul C Roberts, PhD, John H Beigel, MD SAVE Program

**TABLE S1. List of COVAIL US Sites**

**Site name** George Washington Vaccine Research Unit, George Washington University

University of Rochester Medical Center

Hope Clinic of the Emory Vaccine Center

Brigham and Women's Hospital, Harvard Medical School

Saint Louis University, Center for Vaccine Development,

Baylor College of Medicine

University of California San Diego

Emory Center for Childhood Infections and Vaccines of Children’s Healthcare of Atlanta

Duke Human Vaccine Institute, Duke University School of Medicine

University of Illinois at Chicago, Project WISH

University of Texas Medical Branch

Kaiser Permanente Washington Health Research Institute

University of Washington

New York University Manhattan Research Clinic, NYU Grossman School of Medicine

Zuckerberg San Francisco General Hospital, University of California at San Francisco

Morehouse School of Medicine

Washington University School of Medicine

NYU Long Island Research Clinic, NYU Long Island School of Medicine

University of Iowa College of Medicine

Howard University Hospital, Howard University Hospital

University of Alabama at Birmingham

Tulane University School of Medicine

**TABLE S2. Eligible Regimens for Primary Vaccination Series and Boost**

| **Primary series vaccine manufacturer** | **Number of doses in primary series** | **Primary series dose** | **Interval between 1st and 2nd dose** | **Number of booster doses** | **Booster dose** | **Allowed interval between primary series and booster dose*** |
| --- | --- | --- | --- | --- | --- | --- |
| Pfizer BioNTech | 2 | 30 µg | At least 17 days | 1 | 30 µg | ≥ 5 months |
| Moderna | 2 | 100 µg | At least 24 days | 1 | 50 µg | ≥ 5 months |
| Janssen | 1 | 5×10^10^ viral particles | Not applicable | 1 | 5×10^10^ viral particles | ≥ 2 months |

*homologous and heterologous boosters are acceptable

^ no participant had prior NVX-CoV2373 vaccine as booster as this vaccine was approved after the study completed enrollment.

**Table S3: Summary of Demographic and Baseline Characteristics by Vaccination Arm**

|  | **Omicron BA.1  + Wildtype  (N=101)** | **Omicron BA.4/BA.5  + Wildtype  (N=101)** | **Total (N=202)** |
| --- | --- | --- | --- |
| **Age** | | | |
| Median Age, Years (range) | 31 (18-49) | 31 (19-49) | 31 (18-49) |
| Male, No (%) | 45 (45) | 43 (43) | 88 (44) |
| Female, No (%) | 56 (55) | 58 (57) | 114 (56) |
| **Ethnicity, No (%)** | | | |
| Not Hispanic or Latino | 84 (83) | 80 (79) | 164 (81) |
| Hispanic or Latino | 17 (17) | 21 (21) | 38 (19) |
| **Race, No (%)** | | | |
| American Indian or Alaska Native | 1 (1) | 2 (2) | 3 (1) |
| Asian | 11 (11) | 16 (16) | 27 (13) |
| Native Hawaiian or other Pacific Islander | 1 (1) | 0 (0) | 1 (0) |
| Black | 10 (10) | 12 (12) | 22 (11) |
| White | 68 (67) | 65 (64) | 133 (66) |
| Multi Racial | 7 (7) | 4 (4) | 11 (5) |
| Unknown | 3 (3) | 2 (2) | 5 (2) |
| **History of Prior Infection, No(%)** | | | |
| Self-Report | 59 (58) | 60 (59) | 119 (59) |
| Positive Nucleocapsid Protein Antibody | 78 (77) | 72 (71) | 150 (74) |
| Self-Report or Positive Nucleocapsid Protein Antibody | 80 (79) | 76 (75) | 156 (77) |
| **SARS-CoV-2 Vaccination Regimen, No(%)** | | | |
| mRNA Primary, mRNA Boost | 94 (93) | 94 (93) | 188 (93) |
| Ad26 Primary, mRNA Boost | 6 (6) | 6 (6) | 12 (6) |
| Ad26 Primary, Ad26 Boost | 1 (1) | 1 (1) | 2 (1) |
| **Days Since Most Recent Known SARS-CoV-2 Antigenic Exposure, Days, Median (Range)** | | | |
| Most Recent COVID-19 Vaccine | 329.0 (112-454) | 326.0 (112-592) | 326.5 (112-592) |
| Most Recent Self-reported SARS-CoV-2 infection | 252.0 (112-1023) | 278.5 (113-919) | 264.0 (112-1023) |
| Most Recent COVID-19 Vaccine or Self-reported SARS-CoV-2 infection | 288.0 (112-454) | 294.0 (112-585) | 293.0 (112-585) |

**TABLE S4.  Pseudovirus Neutralization Assay Summary Results against variants of concern at baseline, 15 and 29 days after vaccination with Pfizer/BioNTech BNT162b2 Wildtype/Omicron BA.1 for all participants with and without a history a prior infection**

| **Time Point** | **Statistic** | **D614G** | **BA.1** | **BA.4/BA.5** | **B.1.351** | **B.1.617.2** |
| --- | --- | --- | --- | --- | --- | --- |
| Day 1 | n | 99 | 99 | 99 | 99 | 99 |
|  | GMT (95% CI) | 7350 (5886, 9179) | 1080 (824, 1416) | 754 (554, 1025) | 2767 (2125, 3603) | 3277 (2611, 4115) |
|  | Median (Min, Max) | 6756 (193,70071) | 1278 (20,13125) | 946 (20,10852) | 3211 (20,40022) | 3527 (20,24631) |
|  | GMR _D614G_ (95% CI) | 1.0 (NE) | 6.8 (6.0, 7.8) | 9.8 (8.2, 11.6) | 2.7 (2.4, 2.9) | 2.2 (2.1, 2.4) |
|  | Seropositive (95% CI) | 1.00 (0.96, 1.00) | 0.96 (0.90, 0.99) | 0.92 (0.85, 0.96) | 0.99 (0.95, 1.00) | 0.99 (0.95, 1.00) |
| Day 15 | n | 98 | 98 | 98 | 98 | 98 |
|  | GMT (95% CI) | 27000 (23339, 31235) | 6506 (5190, 8155) | 3546 (2860, 4396) | 15183 (12630, 18252) | 14362 (12287, 16788) |
|  | Median (Min, Max) | 26838 (2907,127966) | 6750 (322,80104) | 3491 (320,40673) | 16023 (839,69910) | 14637 (1238,80975) |
|  | GMR _D614G_ (95% CI) | 1.0 (NE) | 4.1 (3.6, 4.8) | 7.6 (6.7, 8.7) | 1.8 (1.6, 1.9) | 1.9 (1.8, 2.0) |
|  | GMFR (95% CI) | 3.6 (3.1, 4.3) | 5.9 (4.9, 7.3) | 4.5 (3.7, 5.6) | 5.4 (4.4, 6.6) | 4.4 (3.7, 5.2) |
|  | Seropositive (95% CI) | 1.00 (0.96, 1.00) | 1.00 (0.96, 1.00) | 1.00 (0.96, 1.00) | 1.00 (0.96, 1.00) | 1.00 (0.96, 1.00) |
| Day 29 | n | 95 | 95 | 95 | 95 | 95 |
|  | GMT (95% CI) | 29924 (25450, 35184) | 4964 (4005, 6152) | 2968 (2396, 3676) | 12816 (10316, 15922) | 12782 (10734, 15222) |
|  | Median (Min, Max) | 30050 (2905,221581) | 5326 (194,43726) | 3080 (207,36167) | 14477 (554,180219) | 12362 (1059,104936) |
|  | GMR _D614G_ (95% CI) | 1.0 (NE) | 6.0 (5.3, 6.9) | 10.1 (8.9, 11.5) | 2.3 (2.1, 2.6) | 2.3 (2.2, 2.5) |
|  | GMFR (95% CI) | 4.1 (3.5, 4.8) | 4.7 (3.9, 5.6) | 3.9 (3.1, 4.7) | 4.7 (3.9, 5.7) | 3.9 (3.3, 4.6) |
|  | Seropositive (95% CI) | 1.00 (0.96, 1.00) | 1.00 (0.96, 1.00) | 1.00 (0.96, 1.00) | 1.00 (0.96, 1.00) | 1.00 (0.96, 1.00) |
| n=Number of subjects with results available at time point  Confidence intervals of the geometric means were calculated with the Student’s t distribution on log-transformed data | | | | | | |

**TABLE S5. Pseudovirus Neutralization Assay Summary Results for variants of concern at baseline, 15 and 29 days after vaccination with Pfizer/BioNTech BNT162b2 Wildtype/Omicron BA.4/BA.5 for participants with and without a history of prior infection**

| **Time Point** | **Statistic** | **D614G** | **BA.1** | **BA.4/BA.5** | **B.1.351** | **B.1.617.2** |
| --- | --- | --- | --- | --- | --- | --- |
| Day 1 | n | 99 | 99 | 99 | 99 | 99 |
|  | GMT (95% CI) | 7735 (6003, 9967) | 994 (719, 1374) | 722 (524, 995) | 2853 (2124, 3832) | 3605 (2814, 4618) |
|  | Median (Min, Max) | 7739 (20,106523) | 1391 (20,16839) | 966 (20,18069) | 3267 (20,53291) | 3930 (20,37042) |
|  | GMR _D614G_ (95% CI) | 1.0 (NE) | 7.8 (6.4, 9.5) | 10.7 (8.8, 13.0) | 2.7 (2.4, 3.1) | 2.1 (2.0, 2.3) |
|  | Seropositive (95% CI) | 0.99 (0.95, 1.00) | 0.92 (0.85, 0.96) | 0.90 (0.82, 0.95) | 0.98 (0.93, 1.00) | 0.99 (0.95, 1.00) |
| Day 15 | n | 98 | 98 | 98 | 98 | 98 |
|  | GMT (95% CI) | 34109 (29493, 39448) | 6603 (5164, 8442) | 5939 (4622, 7631) | 19265 (16023, 23164) | 18332 (15669, 21447) |
|  | Median (Min, Max) | 34651 (2156,311256) | 7099 (20,167899) | 7003 (20,229463) | 20076 (863,226284) | 19417 (1170,170506) |
|  | GMR _D614G_ (95% CI) | 1.0 (NE) | 5.2 (4.3, 6.2) | 5.7 (4.8, 6.9) | 1.8 (1.6, 1.9) | 1.9 (1.8, 1.9) |
|  | GMFR (95% CI) | 4.4 (3.5, 5.5) | 6.6 (5.2, 8.3) | 8.1 (6.3, 10.4) | 6.6 (5.2, 8.5) | 5.0 (4.0, 6.3) |
|  | Seropositive (95% CI) | 1.00 (0.96, 1.00) | 0.99 (0.94, 1.00) | 0.99 (0.94, 1.00) | 1.00 (0.96, 1.00) | 1.00 (0.96, 1.00) |
| Day 29 | n | 93 | 93 | 93 | 93 | 93 |
|  | GMT (95% CI) | 36056 (30554, 42550) | 5341 (4229, 6745) | 5128 (4084, 6440) | 16066 (13107, 19693) | 16136 (13453, 19354) |
|  | Median (Min, Max) | 36088 (2949,378373) | 5423 (237,211797) | 5717 (207,69902) | 18269 (645,181095) | 17510 (909,138136) |
|  | GMR _D614G_ (95% CI) | 1.0 (NE) | 6.8 (5.8, 7.9) | 7.0 (6.1, 8.1) | 2.2 (2.0, 2.5) | 2.2 (2.1, 2.4) |
|  | GMFR (95% CI) | 4.4 (3.7, 5.3) | 5.3 (4.2, 6.7) | 6.9 (5.4, 8.7) | 5.4 (4.4, 6.7) | 4.2 (3.5, 5.1) |
|  | Seropositive (95% CI) | 1.00 (0.96, 1.00) | 1.00 (0.96, 1.00) | 1.00 (0.96, 1.00) | 1.00 (0.96, 1.00) | 1.00 (0.96, 1.00) |
| n=Number of subjects with results available at time point  Confidence intervals of the geometric means were calculated with the Student’s t distribution on log-transformed data | | | | | | |

**TABLE S6. Pseudovirus Neutralization Assay Summary Results for variants of concern at baseline, 15 and 29 days after vaccination with Pfizer/BioNTech BNT162b2 Wildtype/Omicron BA.1 for participants without a history of prior infection**

| **Time Point** | **Statistic** | **D614G** | **BA.1** | **BA.4/BA.5** | **B.1.351** | **B.1.617.2** |
| --- | --- | --- | --- | --- | --- | --- |
| Day 1 | n | 21 | 21 | 21 | 21 | 21 |
|  | GMT (95% CI) | 2806 (1461, 5388) | 304 (135, 688) | 126 (58, 274) | 760 (368, 1569) | 1101 (550, 2207) |
|  | Median (Min, Max) | 2598 (193,33590) | 330 (20,13125) | 144 (20,4895) | 960 (20,15664) | 1224 (20,15850) |
|  | GMR _D614G_ (95% CI) | 1.0 (NE) | 9.2 (6.5, 13.0) | 22.3 (13.5, 36.8) | 3.7 (2.9, 4.6) | 2.5 (2.1, 3.1) |
|  | Seropositive (95% CI) | 1.00 (0.84, 1.00) | 0.81 (0.58, 0.95) | 0.62 (0.38, 0.82) | 0.95 (0.76, 1.00) | 0.95 (0.76, 1.00) |
| Day 15 | n | 20 | 20 | 20 | 20 | 20 |
|  | GMT (95% CI) | 17623 (11946, 25997) | 2846 (1590, 5095) | 1247 (774, 2009) | 7365 (4677, 11597) | 8528 (5629, 12919) |
|  | Median (Min, Max) | 20096 (2907,67475) | 2524 (322,61063) | 1126 (320,15948) | 6557 (839,57509) | 10786 (1238,37368) |
|  | GMR _D614G_ (95% CI) | 1.0 (NE) | 6.2 (4.4, 8.6) | 14.1 (10.7, 18.7) | 2.4 (1.9, 2.9) | 2.1 (1.9, 2.3) |
|  | GMFR (95% CI) | 6.3 (3.8, 10.4) | 9.2 (5.1, 16.5) | 9.0 (5.0, 16.3) | 9.9 (5.7, 17.1) | 7.8 (4.7, 13.0) |
|  | Seropositive (95% CI) | 1.00 (0.83, 1.00) | 1.00 (0.83, 1.00) | 1.00 (0.83, 1.00) | 1.00 (0.83, 1.00) | 1.00 (0.83, 1.00) |
| Day 29 | n | 19 | 19 | 19 | 19 | 19 |
|  | GMT (95% CI) | 16327 (10390, 25655) | 2021 (1158, 3529) | 991 (655, 1500) | 5217 (3154, 8630) | 6459 (4292, 9719) |
|  | Median (Min, Max) | 23743 (2905,66265) | 3058 (194,10102) | 1349 (207,3698) | 6398 (554,27369) | 9093 (1059,20327) |
|  | GMR _D614G_ (95% CI) | 1.0 (NE) | 8.1 (6.4, 10.2) | 16.5 (13.3, 20.4) | 3.1 (2.6, 3.7) | 2.5 (2.2, 2.8) |
|  | GMFR (95% CI) | 6.6 (3.9, 11.3) | 7.9 (4.6, 13.6) | 8.7 (4.9, 15.2) | 8.2 (4.7, 14.2) | 6.8 (4.0, 11.5) |
|  | Seropositive (95% CI) | 1.00 (0.82, 1.00) | 1.00 (0.82, 1.00) | 1.00 (0.82, 1.00) | 1.00 (0.82, 1.00) | 1.00 (0.82, 1.00) |
| n=Number of subjects with results available at time point  Confidence intervals of the geometric means were calculated with the Student’s t distribution on log-transformed data | | | | | | |

**TABLE S7. Pseudovirus Neutralization Assay Summary Results for variants of concern at baseline, 15 and 29 days after vaccination with Pfizer/BioNTech BNT162b2 Wildtype/Omicron BA.4/BA.5 for participants without a history of prior infection**

| **Time Point** | **Statistic** | **D614G** | **BA.1** | **BA.4/BA.5** | **B.1.351** | **B.1.617.2** |
| --- | --- | --- | --- | --- | --- | --- |
| Day 1 | n | 25 | 25 | 25 | 25 | 25 |
|  | GMT (95% CI) | 2452 (1317, 4564) | 163 (80, 333) | 122 (58, 254) | 637 (327, 1237) | 1101 (617, 1963) |
|  | Median (Min, Max) | 3126 (20,32207) | 180 (20,3515) | 139 (20,4186) | 514 (20,9136) | 905 (20,18171) |
|  | GMR _D614G_ (95% CI) | 1.0 (NE) | 15.0 (9.3, 24.3) | 20.1 (11.9, 34.0) | 3.9 (2.8, 5.4) | 2.2 (1.9, 2.6) |
|  | Seropositive (95% CI) | 0.96 (0.80, 1.00) | 0.68 (0.46, 0.85) | 0.60 (0.39, 0.79) | 0.92 (0.74, 0.99) | 0.96 (0.80, 1.00) |
| Day 15 | n | 24 | 24 | 24 | 24 | 24 |
|  | GMT (95% CI) | 26246 (18623, 36990) | 2405 (1304, 4435) | 2198 (1200, 4025) | 11272 (7516, 16905) | 13537 (9427, 19439) |
|  | Median (Min, Max) | 28331 (2156,71209) | 3099 (20,12053) | 2504 (20,14003) | 12184 (863,53266) | 14281 (1170,42697) |
|  | GMR _D614G_ (95% CI) | 1.0 (NE) | 10.9 (6.4, 18.6) | 11.9 (7.0, 20.4) | 2.3 (1.9, 2.8) | 1.9 (1.8, 2.1) |
|  | GMFR (95% CI) | 11.0 (5.8, 20.8) | 15.0 (7.6, 29.5) | 18.1 (8.9, 36.8) | 17.5 (9.1, 33.9) | 12.4 (6.5, 23.5) |
|  | Seropositive (95% CI) | 1.00 (0.86, 1.00) | 0.96 (0.79, 1.00) | 0.96 (0.79, 1.00) | 1.00 (0.86, 1.00) | 1.00 (0.86, 1.00) |
| Day 29 | n | 22 | 22 | 22 | 22 | 22 |
|  | GMT (95% CI) | 25367 (17646, 36468) | 2161 (1389, 3362) | 2151 (1371, 3375) | 8309 (5289, 13051) | 10624 (6994, 16137) |
|  | Median (Min, Max) | 32590 (2949,80338) | 2446 (237,15159) | 1796 (207,18065) | 8368 (645,46619) | 13559 (909,35732) |
|  | GMR _D614G_ (95% CI) | 1.0 (NE) | 11.7 (8.3, 16.5) | 11.8 (8.3, 16.8) | 3.1 (2.4, 3.8) | 2.4 (2.0, 2.9) |
|  | GMFR (95% CI) | 9.3 (5.8, 14.9) | 14.0 (7.1, 27.7) | 18.6 (9.9, 34.7) | 12.4 (6.9, 22.2) | 8.8 (5.6, 14.0) |
|  | Seropositive (95% CI) | 1.00 (0.85, 1.00) | 1.00 (0.85, 1.00) | 1.00 (0.85, 1.00) | 1.00 (0.85, 1.00) | 1.00 (0.85, 1.00) |
| n=Number of subjects with results available at time point  Confidence intervals of the geometric means were calculated with the Student’s t distribution on log-transformed data | | | | | | |

**TABLE S8.  Pseudovirus Neutralization Assay Summary Results against D614G and Omicron Subvariants at baseline and 15 days after vaccination with Pfizer/BioNTech BNT162b2 Wildtype/Omicron BA.1 for a subset of participants with and without a history a prior infection**

| **Time Point** | **Statistic** | **D614G** | **BA.1** | **BA.4/BA.5** | **BQ.1.1** | **XBB.1** |
| --- | --- | --- | --- | --- | --- | --- |
| Day 1 | n | 25 | 25 | 25 | 25 | 25 |
|  | GMT (95% CI) | 2275 (1392, 3718) | 1089 (678, 1749) | 654 (344, 1245) | 154 (90, 265) | 39 (23, 68) |
|  | Median (Min, Max) | 2272 (207,20015) | 962 (97,6668) | 673 (32,8241) | 146 (16,1167) | 40 (5,297) |
|  | GMR _D614G_ (95% CI) | 1.0 (NE) | 2.1 (1.6, 2.8) | 3.5 (2.4, 5.2) | 14.8 (10.7, 20.3) | 57.7 (42.3, 77.7) |
|  | Seropositive (95% CI) | 1.00 (0.86, 1.00) | 1.00 (0.86, 1.00) | 1.00 (0.86, 1.00) | 1.00 (0.86, 1.00) | 0.80 (0.59, 0.93) |
| Day 15 | n | 24 | 24 | 24 | 24 | 24 |
|  | GMT (95% CI) | 10529 (6843, 16201) | 6551 (4442, 9662) | 3587 (2072, 6210) | 794 (551, 1143) | 294 (194, 445) |
|  | Median (Min, Max) | 11823 (746,76974) | 5823 (722,33414) | 3946 (248,24941) | 759 (101,4297) | 282 (20,2199) |
|  | GMR _D614G_ (95% CI) | 1.0 (NE) | 1.6 (1.2, 2.1) | 2.9 (2.0, 4.3) | 13.3 (10.4, 16.9) | 35.8 (27.4, 46.9) |
|  | GMFR (95% CI) | 4.4 (3.1, 6.4) | 5.7 (3.8, 8.5) | 4.9 (3.4, 7.1) | 4.7 (3.3, 6.8) | 6.8 (4.5, 10.5) |
|  | Seropositive (95% CI) | 1.00 (0.86, 1.00) | 1.00 (0.86, 1.00) | 1.00 (0.86, 1.00) | 1.00 (0.86, 1.00) | 1.00 (0.86, 1.00) |

n=Number of subjects with results available at time point 
Confidence intervals of the geometric means were calculated with the Student’s t distribution on log-transformed data

**TABLE S9. Pseudovirus Neutralization Assay Summary Results for D614G and Omicron Subvariants at baseline and 15 days after vaccination with Pfizer/BioNTech BNT162b2 Wildtype/Omicron BA.4/BA.5 for a subset of participants with and without a history of prior infection**

| **Time Point** | **Statistic** | **D614G** | **BA.1** | **BA.4/BA.5** | **BQ.1.1** | **XBB.1** |
| --- | --- | --- | --- | --- | --- | --- |
| Day 1 | n | 25 | 25 | 25 | 25 | 25 |
|  | GMT (95% CI) | 1842 (1222, 2776) | 1213 (683, 2154) | 610 (316, 1177) | 148 (88, 249) | 52 (31, 87) |
|  | Median (Min, Max) | 1414 (523,11934) | 1624 (76,11967) | 729 (27,9429) | 140 (5,2103) | 71 (5,543) |
|  | GMR _D614G_ (95% CI) | 1.0 (NE) | 1.5 (1.1, 2.1) | 3.0 (2.0, 4.5) | 12.4 (8.8, 17.5) | 35.5 (25.2, 50.2) |
|  | Seropositive (95% CI) | 1.00 (0.86, 1.00) | 1.00 (0.86, 1.00) | 1.00 (0.86, 1.00) | 0.96 (0.80, 1.00) | 0.88 (0.69, 0.97) |
| Day 15 | n | 25 | 25 | 25 | 25 | 25 |
|  | GMT (95% CI) | 8967 (5911, 13604) | 6921 (4942, 9691) | 6140 (4321, 8725) | 1077 (810, 1433) | 398 (281, 563) |
|  | Median (Min, Max) | 7594 (1124,63821) | 7754 (888,37374) | 4987 (1713,43534) | 1105 (331,3731) | 402 (113,1883) |
|  | GMR _D614G_ (95% CI) | 1.0 (NE) | 1.3 (0.9, 1.8) | 1.46 (0.9, 2.3) | 8.3 (6.0, 11.5) | 22.6 (15.5, 32.8) |
|  | GMFR (95% CI) | 4.9 (3.0, 7.8) | 5.7 (3.6, 9.1) | 10.1 (6.2, 16.4) | 7.3 (4.6, 11.5) | 7.7 (5.0, 11.9) |
|  | Seropositive (95% CI) | 1.00 (0.86, 1.00) | 1.00 (0.86, 1.00) | 1.00 (0.86, 1.00) | 1.00 (0.86, 1.00) | 1.00 (0.86, 1.00) |

n=Number of subjects with results available at time point 
Confidence intervals of the geometric means were calculated with the Student’s t distribution on log-transformed data

**TABLE S10: Number and Percentage of Subjects Experiencing Solicited Events by Symptom, Maximum Severity and**

**Vaccination Group – Any Symptom**

|  | | **Omicron BA.1+Wildtype(Prototype)  (N=101)** | | **Omicron BA.4/BA.5+Wildtype(Prototype)  (N=101)** | | **All Subjects  (N=202)** | |
| --- | --- | --- | --- | --- | --- | --- | --- |
| **Symptom** | **Severity** | **n** | **%** | **n** | **%** | **n** | **%** |
| Any Symptom | None | 6 | 6 | 8 | 8 | 14 | 7 |
|  | Mild | 58 | 57 | 49 | 49 | 107 | 53 |
|  | Moderate | 32 | 32 | 41 | 41 | 73 | 36 |
|  | Severe | 4 | 4 | 3 | 3 | 7 | 3 |
|  | Not Done | 1 | 1 | . | . | 1 | 0 |
| Severity is the maximum severity reported over all solicited symptoms post dosing for each subject.  N=All subjects with any solicited event data recorded in the database. | | | | | | | |

**TABLE S11: Number and Percentage of Subjects Experiencing Solicited Events by Symptom, Maximum Severity and**

**Vaccination Group – Systemic Symptoms**

|  | | Omicron BA.1+Wildtype(Prototype) (N=101) | | Omicron BA.4/BA.5+Wildtype(Prototype) (N=101) | | All Subjects (N=202) | |
| --- | --- | --- | --- | --- | --- | --- | --- |
| Symptom | Severity | n | % | n | % | n | % |
| Any Systemic Symptom | None | 21 | 21 | 19 | 19 | 40 | 20 |
|  | Mild | 50 | 50 | 42 | 42 | 92 | 46 |
|  | Moderate | 25 | 25 | 38 | 38 | 63 | 31 |
|  | Severe | 4 | 4 | 2 | 2 | 6 | 3 |
|  | Not Done | 1 | 1 | . | . | 1 | 0 |
| Temperature | None | 92 | 92 | 89 | 93 | 181 | 92 |
|  | Mild | 5 | 5 | 4 | 4 | 9 | 5 |
|  | Moderate | 2 | 2 | 3 | 3 | 5 | 3 |
|  | Severe | 1 | 1 | . | . | 1 | 1 |
|  | Not Done | . | . | . | . | . | . |
| Feverishness^a^ | None | 78 | 77 | 68 | 67 | 146 | 72 |
|  | Mild | 12 | 12 | 21 | 21 | 33 | 16 |
|  | Moderate | 9 | 9 | 12 | 12 | 21 | 10 |
|  | Severe | 1 | 1 | . | . | 1 | 0 |
|  | Not Done | 1 | 1 | . | . | 1 | 0 |
| Fatigue | None | 36 | 36 | 28 | 28 | 64 | 32 |
|  | Mild | 44 | 44 | 46 | 46 | 90 | 45 |
|  | Moderate | 18 | 18 | 26 | 26 | 44 | 22 |
|  | Severe | 2 | 2 | 1 | 1 | 3 | 1 |
|  | Not Done | 1 | 1 | . | . | 1 | 0 |
| Myalgia | None | 50 | 50 | 44 | 44 | 94 | 47 |
|  | Mild | 34 | 34 | 37 | 37 | 71 | 35 |
|  | Moderate | 15 | 15 | 20 | 20 | 35 | 17 |
|  | Severe | 1 | 1 | . | . | 1 | 0 |
|  | Not Done | 1 | 1 | . | . | 1 | 0 |
| Arthralgia | None | 80 | 79 | 76 | 75 | 156 | 77 |
|  | Mild | 14 | 14 | 19 | 19 | 33 | 16 |
|  | Moderate | 5 | 5 | 6 | 6 | 11 | 5 |
|  | Severe | 1 | 1 | . | . | 1 | 0 |
|  | Not Done | 1 | 1 | . | . | 1 | 0 |
| Headache | None | 56 | 55 | 59 | 58 | 115 | 57 |
|  | Mild | 32 | 32 | 30 | 30 | 62 | 31 |
|  | Moderate | 10 | 10 | 11 | 11 | 21 | 10 |
|  | Severe | 2 | 2 | 1 | 1 | 3 | 1 |
|  | Not Done | 1 | 1 | . | . | 1 | 0 |
| Nausea | None | 85 | 84 | 86 | 85 | 171 | 85 |
|  | Mild | 13 | 13 | 12 | 12 | 25 | 12 |
|  | Moderate | 2 | 2 | 3 | 3 | 5 | 2 |
|  | Severe | . | . | . | . | . | . |
|  | Not Done | 1 | 1 | . | . | 1 | 0 |
| Severity is the maximum severity reported over all solicited symptoms post dosing for each subject. N=All subjects with any solicited event data recorded in the database. ^a^Fever percentages reflect the number of subjects with at least one measurement available in the data system as the denominator. This denominator may differ from other systemic  symptoms, which are solicited in-clinic at the post-dose assessment. | | | | | | | |

**TABLE S12: Number and Percentage of Subjects Experiencing Solicited Events by Symptom, Maximum Severity and**

**Vaccination Group – Local Symptoms**

|  | | Omicron BA.1+Wildtype(Prototype) (N=101) | | Omicron BA.4/BA.5+Wildtype(Prototype) (N=101) | | All Subjects (N=202) | |
| --- | --- | --- | --- | --- | --- | --- | --- |
| Symptom | Severity | n | % | n | % | n | % |
| Any Local Symptom | None | 20 | 20 | 17 | 17 | 37 | 18 |
|  | Mild | 60 | 59 | 68 | 67 | 128 | 63 |
|  | Moderate | 19 | 19 | 15 | 15 | 34 | 17 |
|  | Severe | 1 | 1 | 1 | 1 | 2 | 1 |
|  | Not Done | 1 | 1 | . | . | 1 | 0 |
| Pain | None | 21 | 21 | 19 | 19 | 40 | 20 |
|  | Mild | 62 | 61 | 68 | 67 | 130 | 64 |
|  | Moderate | 17 | 17 | 14 | 14 | 31 | 15 |
|  | Severe | . | . | . | . | . | . |
|  | Not Done | 1 | 1 | . | . | 1 | 0 |
| Induration/Swelling | None | 81 | 80 | 78 | 77 | 159 | 79 |
|  | Mild | 16 | 16 | 20 | 20 | 36 | 18 |
|  | Moderate | 2 | 2 | 3 | 3 | 5 | 2 |
|  | Severe | 1 | 1 | . | . | 1 | 0 |
|  | Not Done | 1 | 1 | . | . | 1 | 0 |
| Erythema/Redness Measurement (mm) | None | 99 | 98 | 100 | 99 | 199 | 99 |
|  | Mild | 1 | 1 | . | . | 1 | 0 |
|  | Moderate | 1 | 1 | 1 | 1 | 2 | 1 |
|  | Severe | . | . | . | . | . | . |
|  | Not Done | . | . | . | . | . | . |
| Induration/Swelling (mm) | None | 95 | 94 | 95 | 94 | 190 | 94 |
|  | Mild | 2 | 2 | 3 | 3 | 5 | 2 |
|  | Moderate | 4 | 4 | 2 | 2 | 6 | 3 |
|  | Severe | . | . | 1 | 1 | 1 | 0 |
|  | Not Done | . | . | . | . | . | . |
| Severity is the maximum severity reported over all solicited symptoms post dosing for each subject. N=All subjects with any solicited event data recorded in the database. | | | | | | | |

**Figure S1: Maximum Severity of local Solicited Events.**

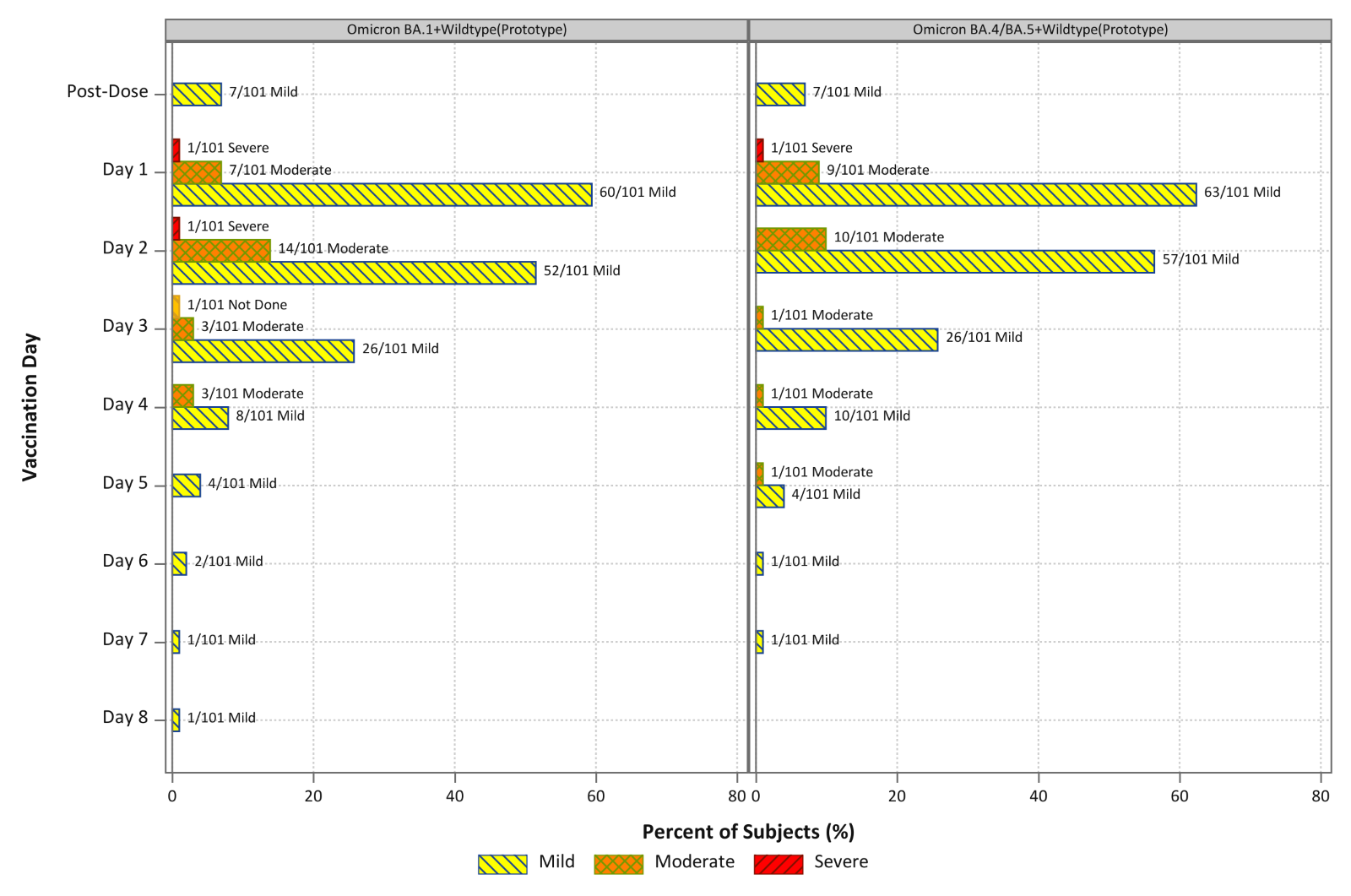

**Figure S2: Maximum Severity of systemic Solicited Events**

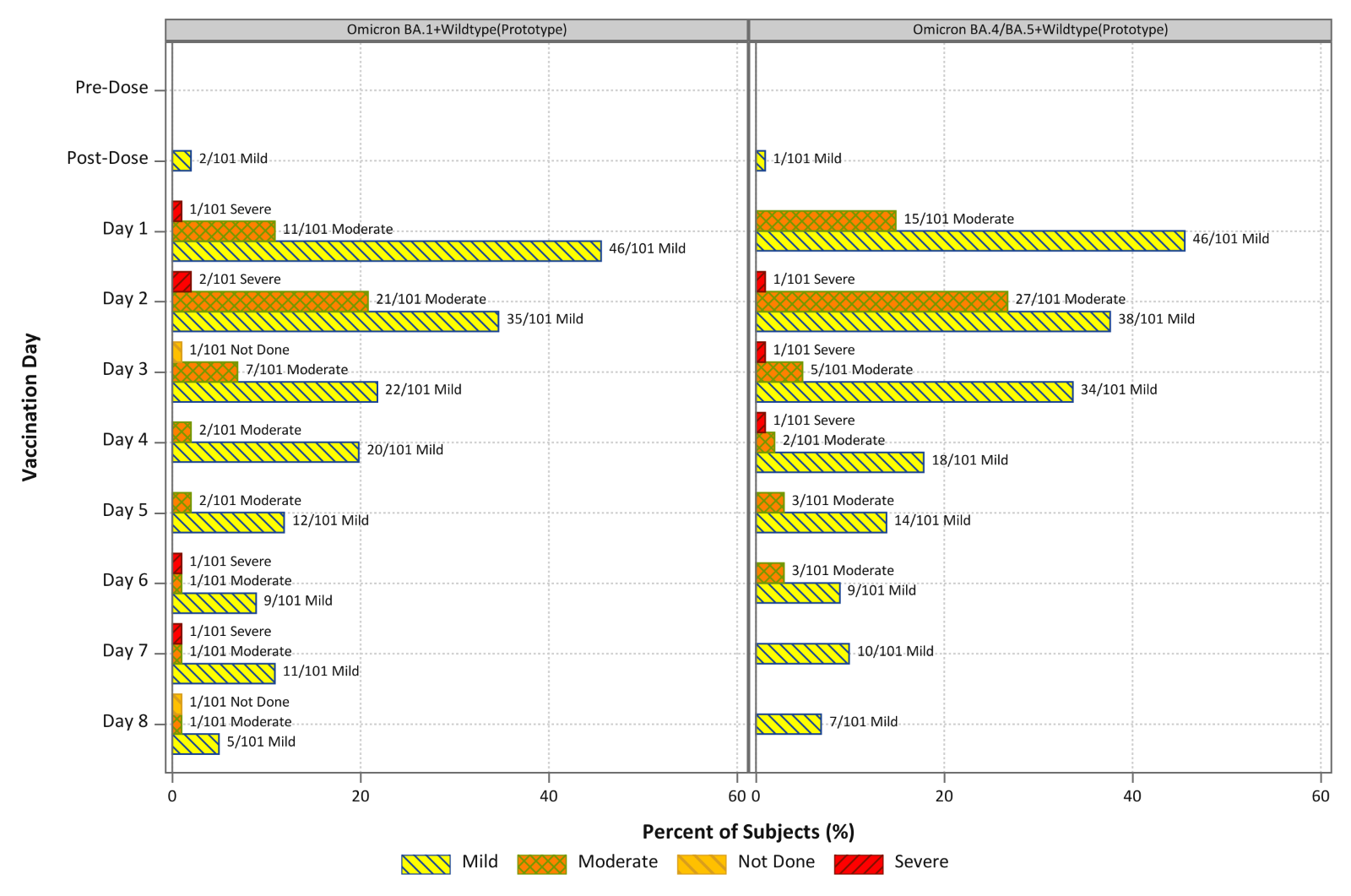
